# Use of antihypertensive drugs and breast cancer risk: a two-sample Mendelian randomization study

**DOI:** 10.1101/2022.05.09.22274758

**Authors:** Guoqiao Zheng, Subhayan Chattopadhyay, Jan Sundquist, Kristina Sundquist, Jianguang Ji

## Abstract

**Background:** Observational studies regarding the correlation between the use of antihypertensive medication and the risk of breast cancer (BC) reported inconsistent findings. We performed a two-sample Mendelian randomization using instrumental variables to proxy changes in gene expressions of antihypertensive medication targets to interrogate this.

**Methods:** We assessed the association between single-nucleotide polymorphisms (SNPs) and drug targetable gene expression with expression quantitative trait loci in blood. Further, we investigated association between the SNPs and BC risk with genome-wide association study summary statistics. We then confirmed the hits from Mendelian randomization with tissue-specific analyses along with additional sensitivity assessments (horizontal pleiotropy, colocalization, multiple tissue enrichment etc.).

**Results:** The overall BC risk was decreased 16% with one standard deviation (SD) increase of *SLC12A2* gene expression in blood (odds ratio, 0.86, 95% confidential interval, 0.78-0.94). This signal was further confirmed in estrogen receptor positive (ER+) BC (0.85, 0.78-0.94). In addition, one SD increase in expression of *PDE1B* in blood was associated with 7% increased risk of ER+ BC (1.07, 1.03-1.11). We detected no evidence of horizontal pleiotropy for these associations and the probability of the causal variants being shared between the gene expression and BC risk was 81.5%, 40.5% and 66.8%, respectively. We failed to observe any significant association between other targeted genes and BC risk.

**Conclusions:** Use of antihypertensive medications that target *SLC12A2* and *PDE1B* is associated with increased and decreased BC risk, respectively.

**Funding:** This work was supported by the Swedish Research Council [2018-02400 to K.S., 2020-01175 to J.S., 2021-01187 to J.J.], Cancerfonden [2017 CAN2017/340 to J.J.], Crafoordska Stiftelsen [to J.J.], MAS Cancer [to J.J.], ALF funding from Region Skåne [to J.J. and K.S.]. The funding body was not involved in the design of the study and collection, analysis, and interpretation of data and in writing the manuscript.

## INTRODUCTION

Nearly one in three adults is estimated to have hypertension globally (1). Antihypertensive drugs are the most commonly prescribed medication in Sweden with over 2.1 million patients taking anti-hypertensive drugs in 2020 (2). These drugs are usually prescribed to hypertensive patients as a long-term management (3). Given the large disease burden and high consumption of the antihypertensive medication, there are concerns surrounding their carcinogenic potential as well as interests regarding their possible anti-cancer effect since the targets of these drugs are widely distributed in normal tissue and may be involved in tumor development. For example, renin-angiotensin system, the target for angiotensin-converting enzyme inhibitors (ACEi) and angiotensin receptor blockers (ARBs), contains components that promote or inhibit cellular proliferation (4). Many observational studies with cohort or case-control design have evaluated the possible link between antihypertensive medication and risk of breast cancer (BC) (5-7), the most common type of cancer and the leading cause of cancer death among women worldwide (8). However, the results are conflicting, as these studies suffer from various biases, including unmeasured confounding, immortal time bias, indication and surveillance bias, which might result in inconsistent conclusions. Due to such shortcomings of the observational studies, previous results could not draw causative conclusions. In addition, there are ethical constraints for conducting randomized clinical trials (RCTs). Therefore, we opted for an alternative design to investigate the causal effect of anti-hypertensive medications on BC risk.

Mendelian randomization (MR) mimics a natural experiment by using genetic variants as a proxy (instrumental variable) for the modifiable exposure (9). The principle is that randomization occurs naturally at conception when genetic variants are allocated at random to individuals from their parents. Consequently, the inherited genetic variants are independent of potential confounding environmental exposures. MR can be regarded as analogous to an RCT that uses genetic variation as the method of randomization ultimately providing causal insights. In addition, the risk estimated from MR reflects a lifetime risk, which is longer than the follow-up in RCT. MR has successfully identified unintended drug effects including adverse drug effects and drug repurposing (10, 11). As for the association between antihypertensive medication and cancer risk, *Yarmolinsky et. al*. reported long-term ACE inhibition to be associated with an increased risk of colorectal cancer by using a MR analysis (12). However, the ARB, a common antihypertensive medication (13), was not included in the analysis. Additionally, the instrumental variable for each class of drug was selected based on only one targeted gene. However, antihypertensive drugs may act on multiple targets involved in different pathways, thus capturing all the possible targets with corresponding instrumental variables for each drug are needed. Finally, the expression quantitative trait loci (eQTL) for the targeted genes from *Yarmolinsky et. al*. was derived only from the whole blood, whereas the targeted genes may be differentially expressed in a tissue-specific manner (14).

Here we aimed to investigate the effect of antihypertensive medication use on BC risk using a two-sample MR design with a consideration of all the commonly prescribed medication for hypertension, and exploring the eQTL from both the whole blood and several other tissues. This study will add evidence to the current knowledge derived from observational studies and try to draw causal conclusions regarding the potential association between antihypertensive medication use on BC risk.

## MATERIAL AND METHODS

This study is based on a couple of publicly available databases, which are summarized in the **Supplementary Table 1**. We used the summary statistics of these publically available databases, and we did not access individual information. We constructed a two-sample MR design where the summary statistics for the association measures are independently evaluated for the exposure (i.e., genetic component and the gene expression) and the outcome (i.e., genetic component and BC). We began by assessing the association between a single-nucleotide polymorphism (SNP) and drug targeted gene expression derived from publicly derived eQTL data in whole blood. In this, three basic assumptions must have held (9): 1) The genetic variants are associated with the risk factor of interest (relevance assumption); 2) There are no unmeasured confounders of the associations between genetic variants and outcome (independence assumption); 3) The genetic variants affect the outcome only through their effect on the risk factor of interest (exclusion restriction). To evaluate and establish the significant MR associations, we further performed tissue-specific analyses to confirm the hits along with additional sensitivity analyses, including assessment of horizontal pleiotropy, colocalization analysis, and multiple tissue enrichment evaluations. The design flow chart is demonstrated in **Figure 1**.

**Figure 1.**
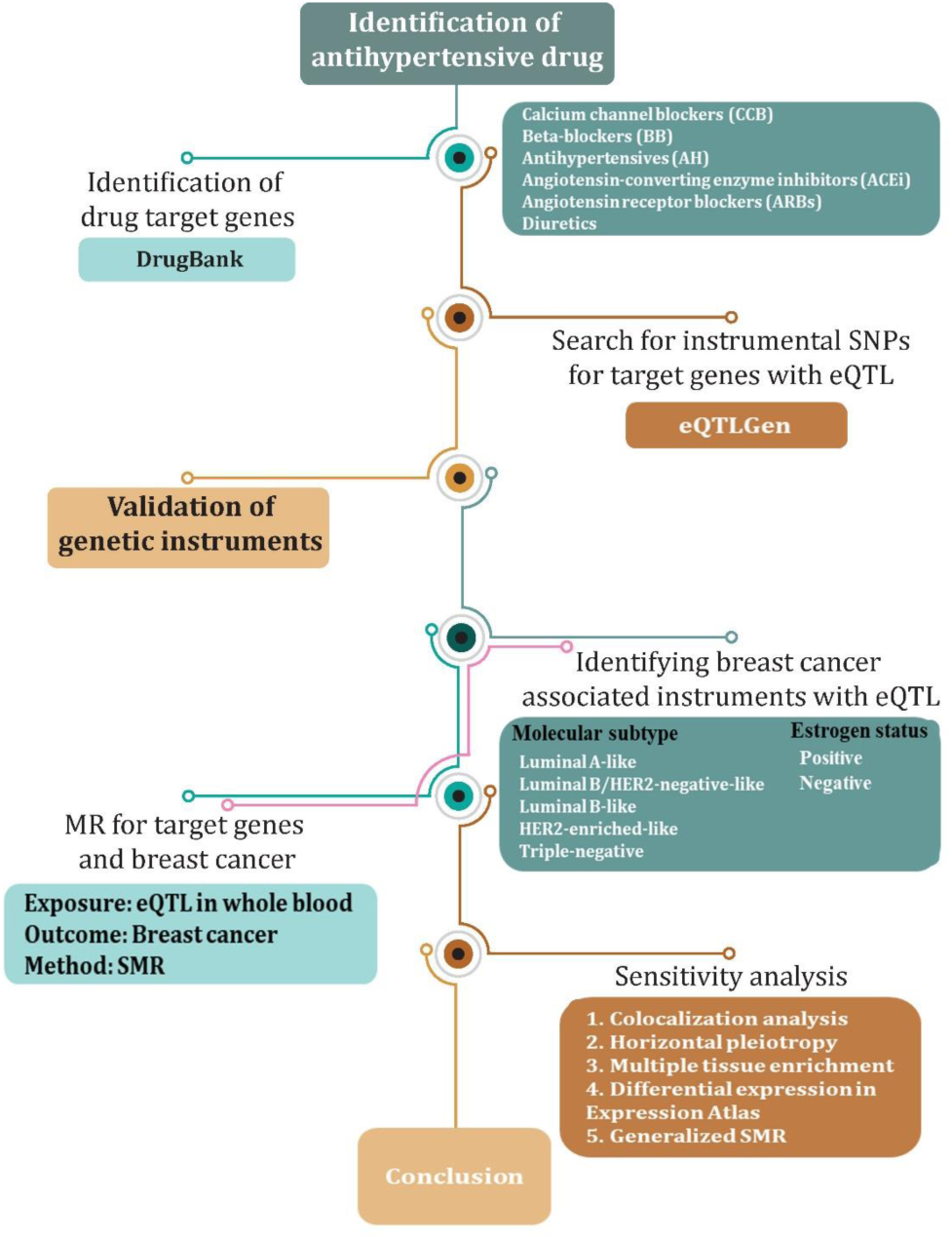
The flowchart of the study. Mendelian randomization, SMR, summary-based MR, eQTL, expression quantitative trait loci, SNP, single-nucleotide polymorphism, HEIDI, heterogeneity in dependent instruments, BC, breast cancer.

### 1. Identification of drug target genes

The commonly prescribed antihypertensive medications, including ACEis, ARBs, beta-blockers (BB), calcium channel blockers (CCB), diuretics and antihypertensives, were considered in the analysis (13). We identified the genes targeted by different classes of anti-hypertensive medication through DrugBank (15).

### 2. Discovery of genetic instruments for targeted gene expression

To identify common SNPs (minor allele frequency >1%) associated with the targeted gene expression of antihypertensive drug in blood, we extracted publicly available expression quantitative trait loci (eQTL) data from eQTLGen (https://www.eqtlgen.org/). The consortium incorporates 37 datasets, with a total of 31,684 individuals and reports on 16,987 genes expressed in whole blood. The eQTL SNPs for this analysis were from *cis*-regulated regions (2 Mb). We used F statistic to assess the strength of the instrumental SNPs.

### 3. Validation of genetic instrument with systolic blood pressure GWAS

Given that we are trying to identify instruments for blood pressure-lowering medication, we performed two-sample MR analysis with gene expression in blood (using the eQTL data) as exposure and systolic blood pressure (SBP) as the outcome to ensure we have valid instruments for the medication of interest. The summary statistics for SBP was a GWAS of SBP in 757 601 individuals of European ancestry drawn from UK Biobank (UKB) and the International Consortium of Blood Pressure Genome Wide Association Studies (ICBP) (16). The summary-based MR (SMR) method (version 1.02) was used to perform the two-sample MR analysis (17). It tests a top SNP against the eQTL summary. For SNPs that passed the significance threshold for the SMR test (i.e., P < 0.05), additionally we performed the HEIDI (heterogeneity in dependent instruments) test to distinguish pleiotropy from linkage with P-value threshold of 0.01, in which up to the top 20 SNPs by default were used for heterogeneity test with LD pruning (0.05 -0.9) (17).

### 4. Accumulation of genetic summary statistics for breast cancer risk

We obtained publicly available GWAS summary statistics for BC risk from the Breast Cancer Association Consortium (BCAC). The analysis for the overall BC risk included 118,474 cases and 96,201 controls of European ancestry participating in 82 studies (18). For analyses of BC molecular subtypes, 7,325 participants were included for luminal A-like cases, 1,779 for luminal B/HER2-negative-like cases, 1,682 for luminal B-like cases, 718 for HER2-enriched-like, 2,006 for triple-negative cases and 20,815 for controls (18). We also obtained the GWAS summary statistics for estrogen receptor-positive (ER+) and -negative (ER-) BC from BCAC, which contained 69,501 ER+ BC cases, 21,468 ER-BC cases and 105,974 controls (19).

### 5. MR analysis between targeted gene expression in blood and breast cancer risk

We performed MR analysis to estimate the association between target gene expression change (using whole blood eQTL) and BC risk (using GWAS). Bonferroni correction was used to identify significant associations due to multiple testing. For the association that reached corrected significant level, we generated SMR locus plot using the method presented on SMR webpage (https://yanglab.westlake.edu.cn/software/smr/#Overview).

### 6. Sensitivity analyses

#### 6.1 Colocalization analysis

This analysis is to assess if two independent association signals at the same locus, typically generated by two GWAS studies, are consistent with a shared causal variant. In the context of our study, we examined if the drug targeted gene expression and occurrence of BC in statistically significant MR associations shared a common causal variant by applying a Bayesian localization approach (20). As a convention, the posterior probability larger than 0.80 is considered supportive for a common causal variant. The R package ‘coloc’ (v3.1, https://cran.r-project.org/web/packages/coloc/) was used to perform the test.

#### 6.2 Assessment of horizontal pleiotropy

A genetic variant can be associated with more than one gene expression, which is known as horizontal pleiotropy. Horizontal pleiotropy is a violation of the instrumental variable assumption (exclusion restriction) that the genetic instrument is associated with the outcome only via changes in gene expression of the drug target. We tested for horizontal pleiotropy by extracting available associations with all other nearby genes (within a 2Mb window) for each genetic instrument. For the nearby genes showing significant association with the genetic instrument, we performed SMR analysis to test if the expression of these genes was associated with BC risk. For those genes with significant SMR associations, colocalization analysis was performed to estimate the probability that two association signals share the same causal variant.

#### 6.3 Multiple tissue sensitivity

We then evaluated the impact of the actionable targets in several different tissues along with whole blood and breast epithelium. Gene set enrichment analysis was performed with the assorted drug target genes to quantify all biological pathways prone to be affected by the drugs. RNA seq data on different tissues (except prostate and testis) were utilized for this purpose and analyzed with R package ‘TissueEnrich’ (21). The enrichment analysis queries expression modulation with fold changes across different human tissue types. A significant departure was defined at an adjusted fold change in expression of value 2 and the fold change was also tested for statistical significance. For tissues that showed significant fold changes, further MR association between tissue specific eQTL and BC risk were retrieved from summary statistics reported by Barbeira et. al. (22).

#### 6.4 Differential gene expression in Expression Atlas

To verify the associations that have shown causal variant and signaled no horizontal pleiotropy, we retrieved transcriptomic data of the significant genes for tumor tissue (or other tissue from BC patients) and normal tissue (or other tissue from healthy controls) on Expression Atlas (https://www.ebi.ac.uk/gxa/home). Differential gene expressions between tissues were evaluated as fold change using normal tissue (or other tissue from healthy controls) as reference.

#### 6.5 MR analysis to estimate the association of SBP with breast cancer

To determine whether the association between drug targeted gene expression and BC risk is likely to be mediated via changes in blood pressure or whether the association may be driven independently, we estimated the effect size for the association between genetically estimated SBP and BC using the generalized summary data-based MR (GSMR) method (23). The GSMR (implemented in Genome-wide Complex Trait Analysis, version 1.91.7) is an extension of SMR that uses multiple genetic variants associated with the risk factor to test for potential causality. The above-mentioned GWAS summary statistics on SBP (exposure) and overall BC risk (outcome) in European individuals were used for the analysis. As usual, a HEIDI P < 0.01 was used to detect outlying variants. In addition, we performed two-sample MR analysis using “*Two-Sample MR*” R package to check the consistency of the signals from the two different methods.

## RESULTS

### Genetic Instrument Selection and Validation

We identified a total of 164 blood pressure modulatory drugs from the WHO Collaborating Centre for Drug Statistics Methodology (https://www.whocc.no/). In DrugBank database, 124 of them were identified to target a total of 154 genes. There were expectedly several overlaps between drug classes and associated targets (**Figure 2**, details in **Supplementary Table 2**). For example, *SLC12A2* can be targeted by quinethazone, bumetanide and torasemide, and at the same time, torasemide can also target on gene *SLC12A1*. Among the 154 chosen targeted genes, 72 genes were identified to have strong association with the eQTL SNPs in eQTLGen (F statistic > 10, P_eQTL_ < 1e-8), among which, 32 were found to be causally associated with SBP with statistical significance (*P* < 0.05). A total of 23 associations were confirmed by the HEIDI outlier test (p_HEIDI > 0.01), which were further considered in the MR analysis with BC as the outcome (**Supplementary Table 3**). It should be noted that the number of SNPs in HEIDI test for gene P4HA1 was less than five (n=3).

**Figure 2.**
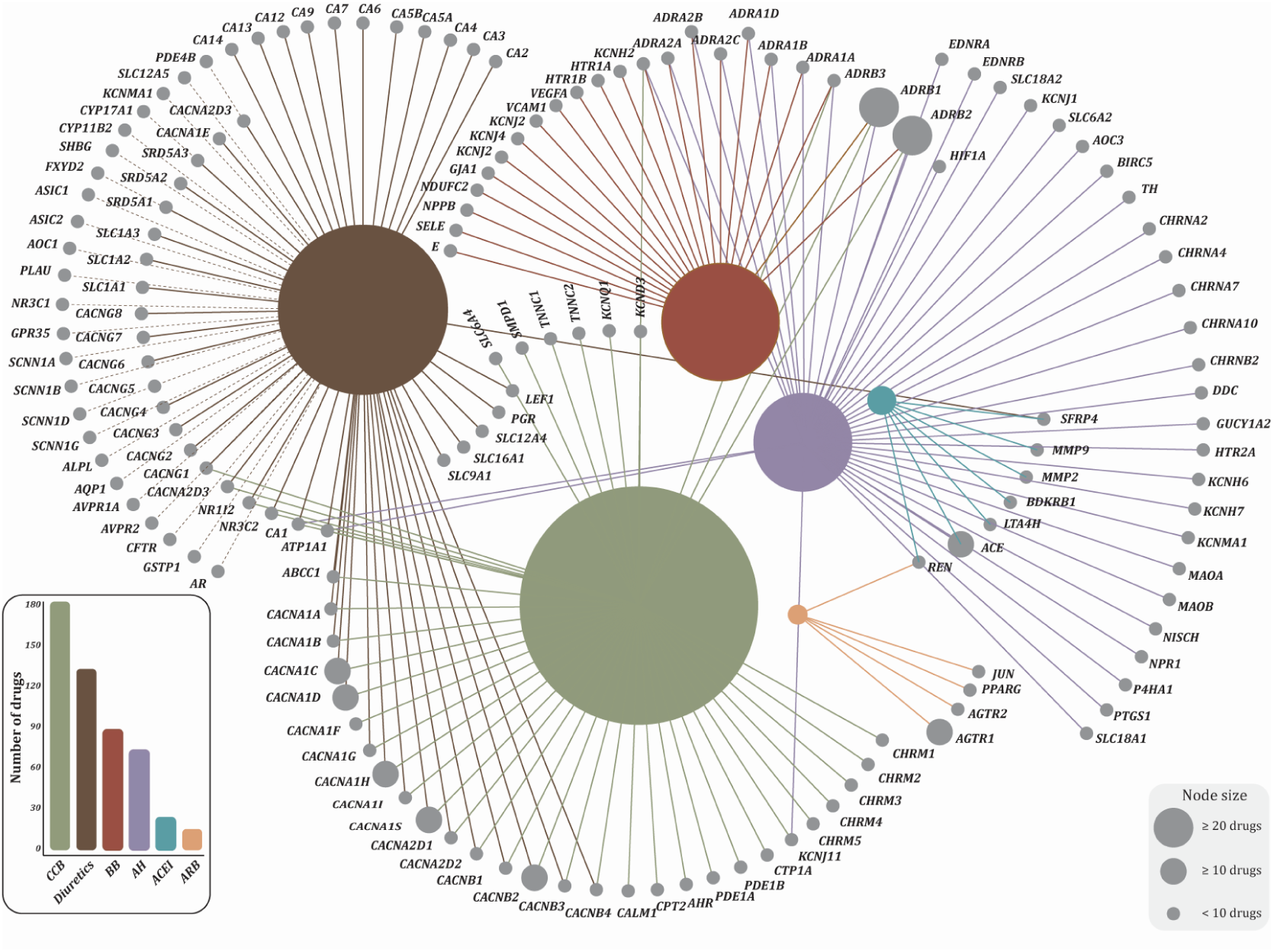
A network of established links between hypertension drugs considered and the corresponding target genes. Each color represents a different class of drugs (annotated in the histogram) and the circle size represents the number of drugs belonging to a class. Abbreviations: CCB, calcium channel blockers, BB, beta-blockers, AH, antihypertensives, ACEi, angiotensin-converting enzyme inhibitors, ARBs, angiotensin receptor blockers.

### MR analysis for association of targeted gene in blood and overall and subtypes BC risk

In the analysis of MR association between the targeted genes in blood and overall BC risk, we identified significant associations for five genes including *P4HA1, SLC12A2, KCNJ11, CA12* and PDE1B, but only the association for SLC12A2 was significant with Bonferroni correction (0.05/23=2.2 × 10^−3^) (**Table 1**). One standard deviation increase in the expression of *SLC12A2* was associated with 14% decrease in BC risk (odds ratio, OR, 0.86, 95% confidence interval, 95%CI, 0.78-0.94). This association was supported by the HEIDI outlier test. The SMR locus plot for this association is shown in **Figure 3**. With the same analysis for ER+ and ER-BC (**Table 2**), a nominal significance was observed in the association between ER+ BC risk and expressions of *P4HA1, SLC12A2, KCNJ11* and *PDE1B*, among which *SLC12A2* and *PDE1B* were still significant after Bonferroni correction (0.05/(23×2)=1.1 × 10^−3^). One SD increase in the *SLC12A2* expression was associated with 15% decrease in the risk of ER+ BC (0.85, 0.78-0.94), while one SD increase in *PDE1B* expression was associated with 7% increased risk (1.07, 1.03-1.11). For ER-BC, only *SLC12A2* showed a nominal significance but it was not significant while considering multiple testing. For the molecular subtype of BC (see **Table 2**), we found *P4HA1, SLC12A2, KCNJ11* and *CA12* were associated with luminal A-like BC with nominal significance, *P4HA1, SLC12A2, ATP1A1, NR3C1, CA12, CACNA1D* and *GPR35* for luminal B-like BC, *SLC12A2* and *SCNN1D* for luminal B/HER2-negative-like BC, ACE for HER2-enriched-like BC and AOC1 for triple-negative BC. However, none of them was significant after Bonferroni correction (0.05/(23×5)=4.3 × 10^−4^).

**Table 1.** MR association between drug target gene expression in blood and overall risk of breast cancer

| Gene | ProbeChr | Probe_bp | topSNP | topSNP_c<br>hr | topSNP_bp | Effect_allele | Other_allele | Freq_Effect_allele | eQTL association |  |  | BC association |  |  | MR association |  |  | HEIDI Test |  |
| --- | --- | --- | --- | --- | --- | --- | --- | --- | --- | --- | --- | --- | --- | --- | --- | --- | --- | --- | --- |
|  |  |  |  |  |  |  |  |  | beta | se | p | beta | se | p | beta | se | p | p | nsnp |
| SLC12A2 | 5 | 127472419 | rs17764730 | 5 | 127357526 | T | C | 0.21 | -0.186 | 0.009 | 3.2E-86 | 0.028 | 0.007 | 7.57E-05 | -0.150 | 0.039 | 1.05E-04 | 1.56E-01 | 20 |
| P4HA1 | 10 | 74811853 | rs6480668 | 10 | 74849326 | G | A | 0.24 | 0.183 | 0.016 | 3.35E-32 | -0.032 | 0.012 | 7.83E-03 | -0.176 | 0.068 | 9.47E-03 | 4.98E-01 | 8 |
| CA12 | 15 | 63643968 | rs12909041 | 15 | 63743677 | A | C | 0.36 | 0.039 | 0.010 | 1.14E-04 | -0.026 | 0.008 | 6.88E-04 | -0.670 | 0.263 | 1.08E-02 | 5.35E-01 | 5 |
| PDE1B | 12 | 54958078 | rs10747699 | 12 | 54955324 | T | C | 0.39 | 0.406 | 0.009 | 0.00E+00 | 0.017 | 0.007 | 1.31E-02 | 0.041 | 0.017 | 1.32E-02 | 3.63E-01 | 3 |
| KCNJ11 | 11 | 17409142 | rs2074310 | 11 | 17421886 | T | C | 0.26 | 0.157 | 0.012 | 4.37E-38 | -0.015 | 0.006 | 1.99E-02 | -0.092 | 0.040 | 2.20E-02 | 8.06E-02 | 20 |
| NR3C1 | 5 | 142736286 | rs4912908 | 5 | 142791133 | G | A | 0.17 | 0.073 | 0.010 | 1.07E-13 | 0.016 | 0.008 | 4.67E-02 | 0.221 | 0.115 | 5.47E-02 | 2.81E-02 | 15 |
| ATP1A1 | 1 | 116934086 | rs6704439 | 1 | 116867616 | A | C | 0.30 | 0.041 | 0.009 | 4.39E-06 | 0.015 | 0.008 | 5.47E-02 | 0.363 | 0.205 | 7.64E-02 | 8.28E-01 | 20 |
| KCNJ2 | 17 | 68170501 | rs9890133 | 17 | 68169005 | G | A | 0.15 | -0.256 | 0.012 | 8.02E-94 | -0.015 | 0.010 | 1.12E-01 | 0.060 | 0.038 | 1.13E-01 | 9.60E-01 | 20 |
| SLC12A1 | 15 | 48540068 | rs964611 | 15 | 48597514 | A | C | 0.15 | 1.664 | 0.009 | 0.00E+00 | -0.012 | 0.009 | 1.78E-01 | -0.007 | 0.005 | 1.78E-01 | 9.86E-01 | 20 |
| CACNA2D2 | 3 | 50470954 | rs62260815 | 3 | 50474624 | A | G | 0.05 | -0.142 | 0.013 | 8.98E-29 | -0.013 | 0.010 | 2.03E-01 | 0.089 | 0.070 | 2.06E-01 | 3.88E-01 | 20 |
| AHR | 7 | 17362011 | rs17643734 | 7 | 17162882 | G | A | 0.03 | 0.478 | 0.016 | 3.7E-184 | 0.015 | 0.013 | 2.70E-01 | 0.030 | 0.028 | 2.71E-01 | 9.83E-01 | 20 |
| ACE | 17 | 61576813 | rs4277405 | 17 | 61548918 | C | T | 0.35 | 0.075 | 0.009 | 1.59E-17 | 0.006 | 0.006 | 3.18E-01 | 0.083 | 0.084 | 3.21E-01 | 9.97E-01 | 20 |
| AQP1 | 7 | 30929070 | rs2075574 | 7 | 30950744 | T | C | 0.37 | 0.113 | 0.013 | 3.77E-19 | -0.008 | 0.009 | 3.50E-01 | -0.073 | 0.078 | 3.53E-01 | 7.67E-01 | 5 |
| SLC9A1 | 1 | 27459389 | rs6683016 | 1 | 27493194 | T | C | 0.28 | -0.096 | 0.008 | 9E-31 | -0.004 | 0.006 | 5.37E-01 | 0.041 | 0.066 | 5.38E-01 | 7.61E-02 | 20 |
| AOC1 | 7 | 150540153 | rs7806458 | 7 | 150476888 | G | A | 0.28 | 0.669 | 0.008 | 0.00E+00 | -0.003 | 0.006 | 6.20E-01 | -0.005 | 0.009 | 6.20E-01 | 7.63E-01 | 20 |
| CACNA1H | 16 | 1237506 | rs117177120 | 16 | 1238875 | G | T | 0.02 | 0.372 | 0.024 | 4.41E-54 | 0.004 | 0.016 | 7.82E-01 | 0.012 | 0.042 | 7.82E-01 | 4.21E-01 | 20 |
| SLC16A1 | 1 | 113477052 | rs1049434 | 1 | 113456546 | A | T | 0.32 | 0.125 | 0.008 | 9.29E-54 | 0.002 | 0.006 | 8.02E-01 | 0.013 | 0.050 | 8.02E-01 | 4.06E-01 | 20 |
| GPR35 | 2 | 241557762 | rs2975788 | 2 | 241571858 | G | A | 0.43 | 0.293 | 0.009 | 6.2E-259 | 0.002 | 0.007 | 8.10E-01 | 0.005 | 0.022 | 8.10E-01 | 5.79E-01 | 20 |
| CACNA1D | 3 | 53687586 | rs9830632 | 3 | 53735766 | G | A | 0.29 | -0.085 | 0.009 | 2E-22 | -0.001 | 0.007 | 9.19E-01 | 0.008 | 0.080 | 9.19E-01 | 3.50E-01 | 20 |
| ADRB2 | 5 | 148207176 | rs2082395 | 5 | 148200600 | A | G | 0.35 | 0.126 | 0.008 | 6.31E-56 | -0.001 | 0.006 | 9.37E-01 | -0.004 | 0.050 | 9.37E-01 | 7.18E-01 | 4 |
| CA4 | 17 | 58237778 | rs34820870 | 17 | 58244021 | G | T | 0.16 | -0.462 | 0.021 | 6.1E-110 | 0.001 | 0.016 | 9.50E-01 | -0.002 | 0.036 | 9.50E-01 | 9.89E-01 | 20 |
| JUN | 1 | 59248125 | rs2716140 | 1 | 59472397 | C | A | 0.39 | -0.228 | 0.008 | 3.5E-177 | 0.000 | 0.007 | 9.57E-01 | 0.002 | 0.030 | 9.57E-01 | 9.06E-01 | 20 |
| SCNN1D | 1 | 1221612 | rs6662635 | 1 | 1220425 | G | A | 0.08 | -0.475 | 0.044 | 4.76E-27 | -0.001 | 0.031 | 9.80E-01 | 0.002 | 0.064 | 9.80E-01 | 9.46E-01 | 20 |

Beta of MR association represents the log odds per one standard deviation increase in gene expression. A significant HEIDI p-value (<0.01) indicates that any association between gene expression and outcome may be due to linkage where there are two distinct causal variants in linkage disequilibrium.
MR, Mendelian randomization, SNP, single nuclear polymorphism, eQTL, expression quantitative trait loci, se, standard error, BC, breast cancer, HEIDI, heterogeneity in dependent instruments, nsnp, number of SNPs for HEIDI test.

**Table 2.** MR association between drug target gene expression in blood and risk of breast cancer subtype according to ER status and molecular characteristics

| BC type |  | Gene | Pr<br>ob<br>eC<br>hr | Probe_bp | topSNP | topS<br>NP_<br>chr | topSNP_b<br>p | Eff<br>ect<br>_al<br>lel<br>e | Ot<br>he<br>r_<br>all<br>ele | Fre<br>q_E<br>ffect<br>_all<br>ele | eQTL association |  |  | BC association |  |  | MR association |  |  | HEIDI Test |  |
| --- | --- | --- | --- | --- | --- | --- | --- | --- | --- | --- | --- | --- | --- | --- | --- | --- | --- | --- | --- | --- | --- |
|  |  |  |  |  |  |  |  |  |  |  | beta | se | p | beta | se | p | beta | se | p | p | ns<br>np |
| Estrogen<br>receptor<br>status | Positive | P4HA1 | 10 | 74811853 | rs6480668 | 10 | 74849326 | G | A | 0.24 | 0.183 | 0.016 | 3.35E-32 | -0.045 | 0.015 | 2.49E-03 | -0.247 | 0.084 | 3.30E-03 | 3.72E-01 | 8 |
|  |  | SLC12A2 | 5 | 127472419 | rs17764730 | 5 | 127357526 | T | C | 0.21 | -0.186 | 0.009 | 3.20E-86 | 0.029 | 0.009 | 6.96E-04 | -0.158 | 0.048 | 8.67E-04 | 1.23E-01 | 20 |
|  |  | KCNJ11 | 11 | 17409142 | rs2074310 | 11 | 17421886 | T | C | 0.26 | 0.157 | 0.012 | 4.37E-38 | -0.016 | 0.008 | 4.20E-02 | -0.099 | 0.050 | 4.53E-02 | 4.33E-01 | 20 |
|  |  | PDE1B | 12 | 54958078 | rs1874308 | 12 | 54954356 | A | T | 0.39 | 0.429 | 0.009 | 0.00E+00 | 0.030 | 0.008 | 3.38E-04 | 0.070 | 0.019 | 3.28E-04 | 2.37E-01 | 14 |
|  | Negative | SLC12A2 | 5 | 127472419 | rs17764730 | 5 | 127357526 | T | C | 0.21 | -0.186 | 0.009 | 3.20E-86 | 0.037 | 0.013 | 5.13E-03 | -0.200 | 0.072 | 5.75E-03 | 7.17E-01 | 20 |
| molecular<br>characteri<br>stics | Luminal<br>A-like | P4HA1 | 10 | 74811853 | rs6480668 | 10 | 74849326 | G | A | 0.24 | 0.183 | 0.016 | 3.35E-32 | -0.044 | 0.016 | 6.35E-03 | -0.238 | 0.090 | 7.84E-03 | 9.37E-01 | 8 |
|  |  | PDE1B | 12 | 54958078 | rs1874308 | 12 | 54954356 | A | T | 0.39 | 0.429 | 0.009 | 0.00E+00 | 0.023 | 0.009 | 9.75E-03 | 0.053 | 0.020 | 9.86E-03 | 5.67E-01 | 14 |
|  |  | CA12 | 15 | 63643968 | rs12909041 | 15 | 63743677 | A | C | 0.36 | 0.039 | 0.010 | 1.14E-04 | -0.024 | 0.010 | 1.51E-02 | -0.632 | 0.307 | 3.97E-02 | 5.06E-01 | 6 |
|  |  | SLC12A2 | 5 | 127472419 | rs17764730 | 5 | 127357526 | T | C | 0.21 | -0.186 | 0.009 | 3.20E-86 | 0.019 | 0.009 | 4.35E-02 | -0.101 | 0.050 | 4.46E-02 | 9.57E-01 | 20 |
|  | Luminal B-like | P4HA1 | 10 | 74811853 | rs6480668 | 10 | 74849326 | G | A | 0.24 | 0.183 | 0.016 | 3.35E-32 | -0.093 | 0.037 | 1.11E-02 | -0.506 | 0.204 | 1.31E-02 | 9.98E-01 | 8 |
|  |  | GPR35 | 2 | 241557762 | rs2975788 | 2 | 241571858 | G | A | 0.43 | 0.293 | 0.009 | 6.23E-259 | -0.044 | 0.019 | 2.02E-02 | -0.151 | 0.065 | 2.05E-02 | 1.21E-02 | 20 |
|  |  | SLC12A2 | 5 | 127472419 | rs17764730 | 5 | 127357526 | T | C | 0.21 | -0.186 | 0.009 | 3.20E-86 | 0.044 | 0.020 | 3.24E-02 | -0.236 | 0.111 | 3.34E-02 | 1.73E-02 | 20 |
|  |  | CACNA1D | 3 | 53687586 | rs9830632 | 3 | 53735766 | G | A | 0.29 | -0.085 | 0.009 | 2.00E-22 | 0.042 | 0.020 | 3.17E-02 | -0.499 | 0.238 | 3.59E-02 | 6.17E-01 | 20 |
|  |  | CA12 | 15 | 63643968 | rs12909041 | 15 | 63743677 | A | C | 0.36 | 0.039 | 0.010 | 1.14E-04 | -0.056 | 0.023 | 1.29E-02 | -1.457 | 0.697 | 3.66E-02 | 8.70E-01 | 6 |
|  |  | ATP1A1 | 1 | 116934086 | rs6704439 | 1 | 116867616 | A | C | 0.30 | 0.041 | 0.009 | 4.39E-06 | 0.050 | 0.022 | 2.10E-02 | 1.216 | 0.590 | 3.92E-02 | 9.63E-01 | 20 |
|  |  | NR3C1 | 5 | 142736286 | rs4912908 | 5 | 142791133 | G | A | 0.17 | 0.073 | 0.010 | 1.07E-13 | 0.047 | 0.023 | 3.76E-02 | 0.649 | 0.324 | 4.53E-02 | 3.31E-01 | 14 |
|  | luminal<br>B/HER2-negative-like | SLC12A2 | 5 | 127472419 | rs17764730 | 5 | 127357526 | T | C | 0.21 | -0.186 | 0.009 | 3.20E-86 | 0.054 | 0.017 | 1.82E-03 | -0.292 | 0.095 | 2.08E-03 | 6.53E-01 | 20 |
|  |  | SCNN1D | 1 | 1221612 | rs6662635 | 1 | 1220425 | G | A | 0.08 | -0.475 | 0.044 | 4.76E-27 | -0.206 | 0.080 | 1.03E-02 | 0.434 | 0.174 | 1.26E-02 | 3.35E-01 | 20 |
|  | HER2-enriched | KCNJ11 | 11 | 17409142 | rs2074310 | 11 | 17421886 | T | C | 0.26 | 0.157 | 0.012 | 4.37E-38 | -0.041 | 0.027 | 1.28E-01 | -0.258 | 0.171 | 1.31E-01 | 6.21E-01 | 20 |
|  |  | ACE | 17 | 61576813 | rs4277405 | 17 | 61548918 | C | T | 0.35 | 0.075 | 0.009 | 1.59E-17 | 0.069 | 0.026 | 7.99E-03 | 0.918 | 0.363 | 1.13E-02 | 1.93E-01 | 20 |
|  | Triple-negative | AOC1 | 7 | 150540153 | rs7806458 | 7 | 150476888 | G | A | 0.28 | 0.669 | 0.008 | 0.00E+00 | -0.031 | 0.016 | 4.97E-02 | -0.046 | 0.024 | 4.97E-02 | 9.62E-01 | 20 |

Beta of MR association represents the log odds per one standard deviation increase in gene expression. A significant HEIDI p-value ( $<0.01$ ) indicates that any association between gene expression and outcome may be due to linkage where there are two distinct causal variants in linkage disequilibrium.
MR, Mendelian randomization, SNP, single nuclear polymorphism, eQTL, expression quantitative trait loci, se, standard error, BC, breast cancer, HEIDI, heterogeneity in dependent instruments, nsnp, number of SNPs for HEIDI test.

**Figure 3.**
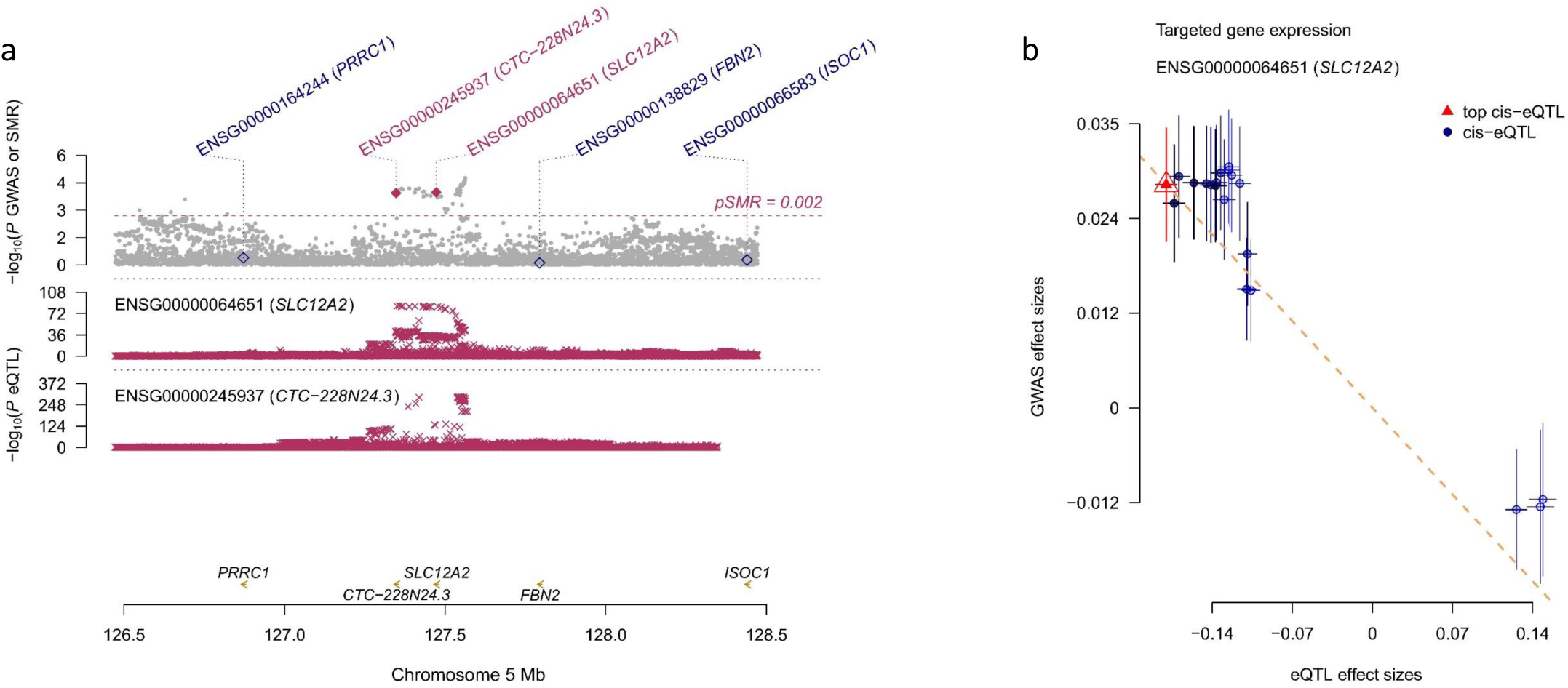
MR association between *SLC12A2* gene expression in blood and breast cancer risk (a) Top, P values from the GWAS for breast cancer (grey dots) and P values from SMR tests (diamonds). Bottom, P values eQTL data for *SLC12A2* and *CTC-228N24*.*3*. Shown in a are all the SNPs available in the GWAS and eQTL data. (b) Effect sizes of the SNPs (used for the HEIDI test) from the GWAS against those from the eQTL data. The orange dashed lines represent the estimate of effect size of the MR association at the top cis-eQTL (rather than the regression line). Error bars are the standard errors of SNP effects.

### Sensitivity analyses

We conducted several sensitivity analyses to evaluate the discovered significant associations between targetable gene expression and BC risk. The associations for the top SNP pertaining to *SLC12A2* and *PDE1B* with nearby genes within the 2MB window are shown in **Supplementary Table 4**. In the test for horizontal pleiotropy, gene *CTC-228N24*.*3* within the 2MB window of *SLC12A2* was also found significant association with BC risk (**Supplementary Table 5**). The P-value of the SMR association for *CTC-228N24*.*3* was similar to that of *SLC12A2*. However, the posterior probability for a common variant between *CTC-228N24*.*3* and overall BC risk was 77%, less than that for *SLC12A2* (81.5%). The posterior probability for a common causal variant between *SLC12A2* expression and risk of ER+ BC was 40.5% and for *PDE1B* was 66.8%. None of the corresponding nearby genes showed a higher probability than *SLC12A2* and *PDE1B*.

In the tissue enrichment analysis, 16 different tissues were found enriched with targeted genes (**Supplementary Figure 1**). We further measured the statistical significance of the associations which largely overlapped with the observation of fold change. Interestingly, smooth muscular, cervical, gallbladder, endometrium, adrenal gland, small seven intestine, placenta, skeletal muscle and kidney were found significant. The MR associations for the available tissue in GTEx are shown in **Supplementary Table 6**. Only expression of *PDE1B* in mammary tissue was negatively associated with BC risk (OR, 0.88, 95%CI, 0.82-0.95), but it was not significant after considering Bonferroni correction (0.05/(23×7)=3.1 × 10^−4^).

We uncovered five data resources in Expression Atlas (see **Supplementary Table 7**), which contained gene expression of *SLC12A2* for invasive BC tissue (or other tissue from BC patients) and normal tissue (or other tissue from healthy controls). The samples were either from blood or breast tissue. Four datasets indicated that *SLC12A2* expression in BC tissue (or blood from BC patients) was lower than that in normal tissue (or blood from healthy controls). Only one showed the opposite association and the type of the BC was invasive ductal carcinoma.

### MR association between systolic blood pressure and overall BC risk

We performed MR with GSMR and TwoSampleMR to test if SBP was associated with overall BC risk using SBP GWAS data as exposure and BC GWAS as the outcome. The results indicated no evidence of the causal association between genetically estimated SBP and BC risk (**Supplementary Table 8**).

### DISCUSSIONS

With this two-sample Mendelian randomization study, we observed that the overall BC risk was decreased 16% along with one SD increase of *SLC12A2* gene expression. *SLC12A2* encodes Na(+)-K(+)-2Cl(-) cotransporter isoform 1 (NKCC1), which can be targeted by loop diuretics including torasemide, bumetanide and quinethazone. Thus, long-term use of these drugs may potentially increase overall BC risk. The significant association with the expression of *SLC12A2* was further observed with ER+ BC. In addition, a SD increase in expression of *PDE1B*, which can be targeted by three CCBs: nicardipine, felodipine and bepridil, was associated with 7% increased risk of ER+ BC. We did not observe any significant association between other targeted genes and BC risk.

A recent meta-analysis evaluated the association between antihypertensive medication use and BC according to 57 conventional observational studies and also reported that use of diuretics was associated with increased BC risk (24). However, this study also found increased BC risk among users of BBs, or CCBs (24), which is inconsistent with our result. Another meta-analysis based on RCTs concluded no consistent evidence that antihypertensive medication use had any effect on cancer risk (25). Our results are in agreement with the findings from *Yarmolinsky et. al*. who investigated antihypertensive drug use and risk of common cancers by evaluating SNPs in *ACE, ADRB1*, and *SLC12A3* in GWAS of SBP to proxy genetic inhibition of the angiotensin-converting enzyme (ACE), β-1 adrenergic receptor (ADRB1), and sodium-chloride symporter (NCC) (12). None of the inhibited targets were associated with the risk of BC (12). Unlike this MR study which only considered one targeted gene for each type of antihypertensive drug, we included all the targets and validated the instruments through systolic blood pressure GWAS.

The mechanisms that links *SLC12A2* gene expression and BC risk are unclear. The GSMR analysis for the association between SBP and BC risk indicates that the effect of the drugs on BC risk was not through blood pressure, which is consistent with the finding that there is no evidence of genetic correlation between blood pressure and BC risk (26). The *SLC12A2*-coded protein, NKCC1 has been reported to be involved in mammary gland development and regulate breast morphogenesis (27). So far, very few studies have investigated the association of BC risk and use of loop diuretics, although among which furosemide targets also NKCC2, a homogeneous protein of NKCC1. A meta-analysis based on four studies found that the relative risk of BC among users of loop diuretics was 0.91(0.77-1.06) compared to nonusers. Elsewhere an in vitro study reported expression of *SLC12A2* in BC cell to be downregulated by the treatment with estrogen (28), which is a well-known risk factor for BC. This finding is in accordance with our current study where *SLC12A2* expression was found protective against BC risk and this association was only significant for ER+ BC. It should be noted that the case number of ER-BC GWAS was much less than ER+ BC GWAS which might detail the loss of signal. Four out of five datasets from Expression Atlas also indicated lower expression of *SLC12A2* in BC tissue (or tissue from BC patients) than the normal tissue (or tissue from controls). On contrary, several in vitro studies reported that inhibition of NKCC1 could reduce cell proliferation, invasion and/or migration in glioblastoma, glioma, esophageal squamous cell carcinoma, hepatocellular carcinoma and gastric cancer cells which may indicate a global underlying tumor-inhibiting mechanism (29-35). Sun et. al. also found that increased expression of NKCC1 was associated with poor prognosis in lung adenocarcinoma and EGFR-mutated adenocarcinoma (36). More studies are required to investigate the role of NKCC1 in BC etiology.

*PDE1B* was the only other gene that showed the association with ER+BC risk. The function of protein PDE1B includes dopaminergic signaling, immune cell activation, and cell survival. It is one of the family members of phosphodiesterases (PDEs) that hydrolyze cyclic adenosine monophosphate (cAMP) and cyclic guanosine monophosphate (cGMP); the latter two participate in many physiological processes such as visual transduction, cell proliferation and differentiation, and cell-cycle regulation (37). Some tumor cells overexpress PDEs and as a consequence, the level of cAMP/cGMP in tumor cells is lower than the normal cells (38).

PDEs have become the potential therapeutic target to increase intracellular cAMP/cGMP and thus inhibit tumor growth (39, 40). PDE1B can be targeted by miR-5701 to inhibit proliferation and promote apoptosis of clear cell renal cell carcinoma cells (41). Wittliff et.al. used gene expression of *PDE1B* and several other genes to predict the overall survival of breast carcinoma (42). However, in the tissue-specific MR, the results for the mammary gland contradict the current evidence, which needs further investigation.

We need to emphasize the strengths and weaknesses of the causal nature of this study design. Use of GWAS summary statistics with two sample MR increases statistical power. The application of genetic variants to proxy the use of antihypertensive medication reduces the chance of confounding, misclassification and immortal time bias that are common caveats in a conventional observational study (43). The different targeted-gene expression was obtained since fetus, which provides the opportunity to observe the effect of antihypertensive medication on BC risk in a long term. Compared to RCT, our study is more efficient as cancer is a late-onset disorder and needs long-term follow-up time. Using the plethora of publicly available summary statistics from GWAS, MR was performed without recruiting new patients or designing additional studies like RCT. We conducted multiple sensitivity analyses such as colocalization analysis and pleiotropy test to evaluate the viability of the assumptions of the instrument variables and to reduce chance findings. As for limitations, the exploration of the molecular subtype of BC was underpowered due to small sample size. Larger GWAS data from the consortium are needed to remedy such issue. The effect of differed gene expression on BC risk due to polymorphism may not be the same as that due to the use of antihypertensive medication, as the former exposure is from early-life time and the latter mostly from adulthood. Therefore, our results provide a strong signal to select existing drugs (NKCC1-targeted antihypertensive medication) to be validated with RCT. Our results may only be applied to European population and research among other population is needed. Additionally, our results suggest future studies could focus on the association between expression level of NKCC1 protein and BC risk, which may provide evidence on risk reduction.

## CONCLUSIONS

By using two-sample MR, we found BC risk was negatively associated with expression of *SLC12A2*, and ER+ BC risk was positively associated with expression of *PDE1B*. Therefore, the use of antihypertensive medication targeting *SLC12A2* and *PDE1B* will be associated respectively increased and decreased risk. The observed effect on the BC risk was independent of systolic blood pressure.

## Data Availability

The data that support the findings of this study are publicly available. The sources have been summarized in Supplementary Table 1. Any request regarding the data from this study should go to Z.G.

## DECLARATIONS

### Availability of data and materials

The data that support the findings of this study are publicly available. The sources have been summarized in **Supplementary Table 1**. Any request regarding the data from this study should go to Z.G..

### Ethical approval and consent to participate

All the data for this study were publicly available summary statistics. Therefore, ethical approval and consent to participate were not required.

### Consent for publication

Not applicable.

### Author contributions

Design: GZ, JJ. Work platform access: JS, KS. Statistical analysis and interpretation: GZ, SC, JJ, JS, KS. Manuscript writing: GZ and all other authors. Approval of the final text: All authors.

