## Supplemental tables for "Use of antihypertensive drugs and breast cancer risk: a two-sample Mendelian randomization study"

**Supplementary Table 1** Data resource used in the current study

| **Category** | **Resource** | **URL** |
|---|---|---|
| List of antihypertensive drugs | WHO Collaborating Centre for Drug Statistics Methodology | https://www.whocc.no/ |
| Identification of targeted gene | Drugbank | https://go.drugbank.com/ |
| Genetic instruments for targeted gene expression in blood | eQTLGen Consortium | https://www.eqtlgen.org/ |
| Systolic blood pressure GWAS | GRASP | https://grasp.nhlbi.nih.gov/FullResults.aspx |
| Breast cancer GWAS | Breast Cancer Association Consortium | https://bcac.ccge.medschl.cam.ac.uk/bcacdata/oncoarray/oncoarray-and-combined-summary-result/ |
| MR association between targeted gene expression in other tissue and risk of breast cancer | Exploiting the GTEx resources to decipher the mechanisms at GWAS loci | https://zenodo.org/record/3942704#.Ya3sANDMKUn |
| Differential gene expression | Expression Atlas | https://www.ebi.ac.uk/gxa/home |

**Supplementary Table 2** Genes targeted by antihypertensive medications in DrugBank

| Drug Class | Drug subclass | Medication Subclass | ATC Code | Drug name | ATC Code | Target Gene(s) |
|---|---|---|---|---|---|---|
| Antihyper tensives | Anti-adrenergic agents, centrally-acting | Rauwolfia alkaloids | C02AA | rescinnamine | C02AA01 | ACE |
| | | | | reserpine | C02AA02 | BIRC5, SLC18A1, SLC18A2 |
| | | | | rauwolfia alkaloids | C02AA04 | SLC18A2 |
| | | | | deserpidine | C02AA05 | SLC18A2 |
| | | | | methoserpidine | C02AA06 | |
| | | | | bietaserpine | C02AA07 | |
| | | Methyldopa | C02AB | methyldopa | C02AB01, C02AB02 | ADRA2A, DDC |
| | | Imidazoline receptor agonists | C02AC | clonidine | C02AC01 | ADRA2A, ADRA2C, ADRA1A, ADRA2B, ADRA1B, ADRA1D |
| | | | | guanfacine | C02AC02 | ADRA2B, ADRA2A |
| | | | | tolonidine | C02AC04 | |
| | | | | moxonidine | C02AC05 | ADRA2A, NISCH |
| | | | | rilmenidine | C02AC06 | ADRA2A |
| | Anti-adrenergic agents, ganglion-blocking | Sulfonium derivatives | C02BA | trimetaphan | C02BA01 | CHRNA10 |
| | | Secondary and tertiary amines | C02BB | mecamylamine | C02BB01 | CHRNA2, CHRNA4, CHRNB2, CHRNA7 |
| | Anti-adrenergic agents, peripherally-acting | Alpha-adrenoreceptor antagonists | C02CA | prazosin | C02CA01 | ADRA1A, ADRA1B, ADRA1D, KCNH2, ADRA2A, ADRA2B |
| | | | | indoramin | C02CA02 | ADRA1A |
| | | | | trimazosin | C02CA03 | |
| | | | | doxazosin | C02CA04 | ADRA1A, ADRA1B, ADRA1D, KCNH2, KCNH6, KCNH7 |
| | | | | urapidil | C02CA06 | |
| | | Guanidine derivatives | C02CC | betanidine | C02CC01 | KCNJ1, ADRA2A, ADRA2B, ADRA2C, ADRB1, ADRB2, ADRB3 |
| | | | | guanethidine | C02CC02 | SLC6A2 |
| | | | | guanoxan | C02CC03 | |
| | | | | debrisoquine | C02CC04 | SLC6A2 |

| | | | | | | |
|---|---|---|---|---|---|---|
| | | | | guanoclor | C02CC05 | |
| | | | | guanazodine | C02CC06 | |
| | | | | guanoxabenz | C02CC07 | |
| | arteriolar smooth muscle, agents acting on | Thiazide derivatives | C02DA | diazoxide | C02DA01 | CA1, CA2, KCNMA1, KCNJ11, ATP1A1 |
| | | Hydrazinophthalazine derivatives | C02DB | dihydralazine | C02DB01 | |
| | | | | hydralazine | C02DB02 | P4HA1, AOC3, HIF1A |
| | | | | endralazine | C02DB03 | |
| | | | | cadralazine | C02DB04 | |
| | | Pyrimidine derivatives | C02DC | minoxidil | C02DC01 | PTGS1, REN, KCNJ1 |
| | | Nitroferricyanide derivatives | C02DD | nitroprusside | C02DD01 | NPR1 |
| | | Guanidine derivatives | C02DG | pinacidil | C02DG01 | |
| | other antihypertensives | Alkaloids, excl. rauwolfia | C02KA | veratrum | C02KA01 | |
| | | Tyrosine hydroxylase inhibitors | C02KB | metirosine | C02KB01 | TH |
| | | MAO inhibitors | C02KC | pargyline | C02KC01 | MAOB, MAOA |
| | | Serotonin antagonists | C02KD | ketanserin | C02KD01 | HTR2A |
| | | Antihypertensives for pulmonary arterial hypertension | C02KX | bosentan | C02KX01 | EDNRB, EDNRA |
| | | | | ambrisentan | C02KX02 | EDNRB, EDNRA |
| | | | | sitaxentan | C02KX03 | EDNRB, EDNRA |
| | | | | macitentan | C02KX04 | EDNRB, EDNRA |
| | | | | riociguat | C02KX05 | GUCY1A2 |
| ACEi | ACE inhibitors, plain | ACE inhibitors, plain | C09AA | captopril | C09AA01 | MMP9, ACE, MMP2, LTA4H, BDKRB1 |
| | | | | enalapril | C09AA02 | ACE |
| | | | | lisinopril | C09AA03 | ACE, REN |
| | | | | perindopril | C09AA04 | ACE, SFRP4 |
| | | | | ramipril | C09AA05 | ACE, BDKRB1 |
| | | | | quinapril | C09AA06 | ACE |
| | | | | benazepril | C09AA07 | ACE |
| | | | | cilazapril | C09AA08 | ACE |

| | | | | | | |
|---|---|---|---|---|---|---|
| | | | | fosinopril | C09AA09 | ACE |
| | | | | trandolapril | C09AA10 | ACE |
| | | | | spirapril | C09AA11 | ACE |
| | | | | delapril | C09AA12 | |
| | | | | moexipril | C09AA13 | ACE |
| | | | | temocapril | C09AA14 | ACE |
| | | | | zofenopril | C09AA15 | ACE |
| | | | | imidapril | C09AA16 | |
| ARBs | Angiotensin ii receptor blockers (ARBs), plain | Angiotensin II receptor blockers (ARBs), plain | C09CA | losartan | C09CA01 | AGTR1 |
| | | | | eprosartan | C09CA02 | AGTR2 |
| | | | | valsartan | C09CA03 | AGTR3 |
| | | | | irbesartan | C09CA04 | AGTR1, JUN |
| | | | | tasosartan | C09CA05 | AGTR1, AGTR2 |
| | | | | candesartan | C09CA06 | AGTR1 |
| | | | | telmisartan | C09CA07 | AGTR1, PPARG |
| | | | | olmesartan medoxomil | C09CA08 | AGTR1 |
| | | | | azilsartan medoxomil | C09CA09 | AGTR2 |
| | | | | fimasartan | C09CA10 | AGTR3 |
| | | Renin-inhibitors | C09XA | remikiren | C09XA01 | REN |
| | | | | aliskiren | C09XA02 | REN |
| CCBs | Selective calcium channel blockers with mainly vascular effects | Dihydropyridine derivatives | C08CA | amlodipine | C08CA01 | CACNA1I, CA1, CACNB1, CACNA1C, SMPD1, CACNA1B, CACNA2D3 |
| | | | | felodipine | C08CA02 | CALM1, CACNA1C, NR3C2, CACNA2D1, CACNA1H, TNNC1, CACNB2, CACNA1S, CACNA1D, CACNA2D2, PDE1B, PDE1A, TNNC2 |
| | | | | isradipine | C08CA03 | CACNA1C, CACNA2D1, CACNA1H, CACNB2, CACNA1S, CACNA1D, CACNA2D2 |

| | | | | | | |
|---|---|---|---|---|---|---|
| | | | | nicardipine | C08CA04 | CHRM3, CHRM1, CHRM5, CHRM4, CALM1, CACNA1C, ADRA1A, CHRM2, ADRA1B, CACNA2D1, ADRA1D, CACNB2, CACNA1D, PDE1B, PDE1A |
| | | | | nifedipine | C08CA05 | CALM1, CACNA1C, NR1I2, CACNB2, CACNA1S, CACNA1D, KCND3, CACNA1G, CACNA1H, CACNA1I |
| | | | | nimodipine | C08CA06 | CACNB1, CACNA1C, NR3C2, CACNB2, CACNA1S, CACNB4, CACNA1F, CACNB3, CACNA1D, AHR |
| | | | | nisoldipine | C08CA07 | CACNA1C, CACNA2D1, CACNB2, CACNA1S, CACNA1D |
| | | | | nitrendipine | C08CA08 | CACNA1C, CACNA2D1, CACNA1H, CACNB2, CACNA1S, CACNA1D, CACNA2D2 |
| | | | | lacidipine | C08CA09 | CACNA1C, CACNA1D, CACNA1F, CACNA1S, CACNB1, CACNB2, CACNB3, CACNB4, CACNA1A |
| | | | | nilvadipine | C08CA10 | CACNA1C, CACNA2D1, CACNB2, CACNA1S, CACNA1D, CACNA2D3 |
| | | | | manidipine | C08CA11 | CACNA1C, CACNA1D, CACNA1F, CACNA1S, CACNB1, CACNB2, CACNB3, CACNB4, CACNA1A, CACNA1G, CACNA1H, CACNA1I |
| | | | | barnidipine | C08CA12 | |
| | | | | lercanidipine | C08CA13 | CACNG1 |
| | | | | cilnidipine | C08CA14 | CACNA1B, CACNA1C, CACNA1D, CACNA1F, CACNA1S, CACNB1, CACNB2, CACNB3, CACNB4, CACNA1A |
| | | | | benidipine | C08CA15 | CACNA1B, CACNA1C, CACNA1D, CACNA1F, CACNA1S, CACNB1, CACNB2, CACNB3, CACNB4, CACNA1A, CACNA1G, CACNA1H, CACNA1I |
| | | | | clevidipine | C08CA16 | CACNA1C, CACNA1S, CACNA1F, CACNA1D |
| | | Other selective calcium channel blockers with mainly vascular effects | C08CX | mibefradil | C08CX01 | CACNA1I, CACNB1, CACNA1C, CACNA1G, CACNA1H, CACNB2, CACNA1S, CACNB4, CACNA1F, CACNB3, CACNA1D |
| | Selective calcium channel blockers with direct cardiac effects | Phenylalkylamine derivatives | C08DA | verapamil | C08DA01 | KCNH2, CACNA1A, CACNA1C, CACNA1G, ADRA1A, ADRA1B, KCNJ11, ADRA1D, SLC6A4, CACNA1H, CACNA1B |
| | | | | gallopamil | C08DA02 | |
| | | Benzothiazepine derivatives | C08DB | diltiazem | C08DB01 | CACNA1C, CACNG1 |
| | | | C08EA | fendiline | C08EA01 | |

| | | | | | | |
|---|---|---|---|---|---|---|
| | Non-selective calcium channel blockers | Phenylalkylamine derivatives | | bepridil | C08EA02 | KCNH2, CACNA1A, CALM1, KCNQ1, ATP1A1, CACNA1H, TNNC1, CACNA2D2, PDE1B, PDE1A |
| | | Other non-selective calcium channel blockers | C08EX | lidoflazine | C08EX01 | |
| | | | | perhexiline | C08EX02 | KCNH2, CPT2, CPT1A |
| Beta blocker | Beta blocking agents | Beta blocking agents, non-selective | C07AA | alprenolol | C07AA01 | ADRB1, HTR1A, ADRB2, ADRB3 |
| | | | | oxprenolol | C07AA02 | ADRB1, ADRB2, ADRB3 |
| | | | | pindolol | C07AA03 | ADRB1, HTR1A, ADRB2, HTR1B, ADRB3 |
| | | | | propranolol | C07AA05 | ADRB1, HTR1A, ADRB2, HTR1B, ADRB3 |
| | | | | timolol | C07AA06 | ADRB1, ADRB2, E |
| | | | | sotalol | C07AA07 | KCNH2, ADRB1, ADRB2 |
| | | | | nadolol | C07AA12 | ADRB1, ADRB2 |
| | | | | mepindolol | C07AA14 | |
| | | | | carteolol | C07AA15 | ADRB1, ADRB2 |
| | | | | tertatolol | C07AA16 | |
| | | | | bopindolol | C07AA17 | ADRB1, HTR1A, ADRB2, HTR1B, ADRB3 |
| | | | | bupranolol | C07AA19 | ADRB1, ADRB2, ADRB3 |
| | | | | penbutolol | C07AA23 | ADRB1, HTR1A, ADRB2, HTR1B |
| | | | | cloranolol | C07AA27 | |
| | Beta blocking agents | Beta blocking agents, selective | C07AB | practolol | C07AB01 | ADRB1 |
| | | | | metoprolol | C07AB02 | ADRB1, ADRB2 |
| | | | | atenolol | C07AB03 | ADRB1, ADRB2 |
| | | | | acebutolol | C07AB04 | ADRB1, ADRB2 |
| | | | | betaxolol | C07AB05 | ADRB1, ADRB3 |
| | | | | bevantolol | C07AB06 | ADRB1, ADRA1A, ADRB2 |
| | | | | bisoprolol | C07AB07 | ADRB1, ADRB2 |
| | | | | celiprolol | C07AB08 | ADRB1, ADRA2A, ADRA2C, ADRA2B, ADRB2, ADRB3 |
| | | | | esmolol | C07AB09 | ADRB1 |
| | | | | epanolol | C07AB10 | |
| | | | | s-atenolol | C07AB11 | |
| | | | | nebivolol | C07AB12 | ADRB1, ADRB2, ADRB3 |
| | | | | talinolol | C07AB13 | |
| | | | | landiolol | C07AB14 | |

| | | | | | | |
|---|---|---|---|---|---|---|
| | Beta blocking agents | Alpha and beta blocking agents | C07AG | labetalol | C07AG01 | ADRB1, ADRB2, ADRA1A, ADRA1B, ADRA1D |
| | | | | carvedilol | C07AG02 | KCNH2, VEGFA, ADRB1, NDUFC2, ADRA2A, ADRA2C, ADRA1A, ADRA2B, ADRA1B, ADRB2, ADRA1D, NPPB, SELE, GJA1, VCAM1, HIF1A, KCNJ4, KCNJ2 |
| Diuretic | Low-ceiling diuretics, thiazides | Thiazides, plain | C03AA | bendroflumethiazide | C03AA01 | CA1, CA2, SLC12A3, CA4, KCNMA1 |
| | | | | hydroflumethiazide | C03AA02 | CA1, CA2, SLC12A1, CA4, KCNMA1, ATP1A1, CA12, CA9, CA7 |
| | | | | hydrochlorothiazide | C03AA03 | SLC12A3, KCNMA1 |
| | | | | chlorothiazide | C03AA04 | CA1, CA2, SLC12A3 |
| | | | | polythiazide | C03AA05 | SLC12A3 |
| | | | | trichlormethiazide | C03AA06 | CA1, CA2, SLC12A3, CA4, ATP1A1 |
| | | | | cyclopenthiazide | C03AA07 | |
| | | | | methyclothiazide | C03AA08 | CA1, CA2, SLC12A1, CA4 |
| | | | | cyclothiazide | C03AA09 | FXYD2, CA1, CA2, SFRP4, CA13, CA14, CA3, CA4, CA5A, CA5B, CA6, CA7, CA9 |
| | | | | mebutizide | C03AA13 | |
| | Low-ceiling diuretics, excl. thiazides | Sulfonamides, plain | C03BA | quinethazone | C03BA02 | CA1, SLC12A2, CA2, SLC12A3, SLC12A1 |
| | | | | clopamide | C03BA03 | |
| | | | | chlortalidone | C03BA04 | CA1, SLC12A1 |
| | | | | mefruside | C03BA05 | |
| | | | | clofenamide | C03BA07 | |
| | | | | metolazone | C03BA08 | SLC12A3 |
| | | | | meticrane | C03BA09 | |
| | | | | xipamide | C03BA10 | |
| | | | | indapamide | C03BA11 | SLC12A3 |
| | | | | clorexolone | C03BA12 | |
| | | | | fenquizone | C03BA13 | |
| | | Mercurial diuretics | C03BC | mersalyl | C03BC01 | SLC16A1, AQP1, ALPL |
| | | Xanthine derivatives | C03BD | theobromine | C03BD01 | ADORA1, PDE4B, ADORA2A |
| | | Other low-ceiling diuretics | C03BX | cicletanine | C03BX03 | |
| | High-ceiling diuretics | Sulfonamides, plain | C03CA | furosemide | C03CA01 | CA2, SLC12A1, GPR35 |
| | | | | bumetanide | C03CA02 | SLC12A5, SLC12A2, SLC12A1, SLC12A4, CFTR |

| | | | | | | |
|---|---|---|---|---|---|---|
| | | | | piretanide | C03CA03 | SLC12A1 |
| | | | | torasemide | C03CA04 | SLC12A2, SLC12A1 |
| | | Aryloxyacetic acid derivatives | C03CC | etacrynic acid | C03CC01 | SLC12A1, ATP1A1, GSTP1, LEF1 |
| | | | | tienilic acid | C03CC02 | |
| | | Pyrazolone derivatives | C03CD | muzolimine | C03CD01 | |
| | | Other high-ceiling diuretics | C03CX | etozolin | C03CX01 | |
| | Potassium-sparing agents | Aldosterone antagonists | C03DA | spironolactone | C03DA01 | AR, CACNA1A, CACNA1B, CACNA1C, CACNA1D, CACNA1E, CACNA1F, CACNA1G, CACNA1H, CACNA1I, CACNA1S, CACNA2D1, CACNA2D2, CACNA2D3, CACNA2D4, CACNB1, CACNB2, CACNB3, CACNB4, CACNG1, CACNG2, CACNG3, CACNG4, CACNG5, CACNG6, CACNG7, CACNG8, CYP11B2, CYP17A1, NR1I2, NR3C1, NR3C2, PGR, SHBG, SRD5A1, SRD5A2, SRD5A3 |
| | | | | potassium canrenoate | C03DA02 | |
| | | | | canrenone | C03DA03 | |
| | | | | eplerenone | C03DA04 | NR3C2 |
| | | Other potassium-sparing agents | C03DB | amiloride | C03DB01 | AOC1, ASIC1, ASIC2, PLAU, SCNN1A, SCNN1B, SCNN1D, SCNN1G, SLC9A1 |
| | | | | triamterene | C03DB02 | SCNN1A, SCNN1B, SCNN1D, SCNN1G |
| | Other diuretics | Vasopressin antagonists | C03XA | tolvaptan | C03XA01 | AVPR1A, AVPR2 |
| | | | | conivaptan | C03XA02 | AVPR1A, AVPR3 |

**Supplementary Table 3** MR association between drug target gene expression in blood and systolic blood pressure

| Gene | Probe Chr | Probe_bp | topSNP | topSNP_chr | topSNP_bp | Effect_allele | Other_allele | Freq_Effect_allele | eQTL association | | | | SBP association | | | MR association | | | HEIDI Test | |
|---|---|---|---|---|---|---|---|---|---|---|---|---|---|---|---|---|---|---|---|---|
| | | | | | | | | | beta | se | p | F_statistics | beta | se | p | beta | se | p | p_HEIDI | nsnp |
| P4HA1 | 10 | 74811853 | rs6480668 | 10 | 74849326 | G | A | 0.07 | 0.183 | 0.016 | 3.35E-32 | 139.5 | 0.304 | 0.061 | 7.17E-07 | 1.655 | 0.362 | 4.82E-06 | 4.41E-02 | 3 |
| ACE | 17 | 61576813 | rs4308 | 17 | 61559625 | A | G | 0.38 | 0.075 | 0.009 | 2.84E-17 | 71.5 | 0.273 | 0.031 | 3.11E-18 | 3.639 | 0.600 | 1.35E-09 | 1.10E-01 | 17 |
| SLC12A2 | 5 | 127472419 | rs17764730 | 5 | 127357526 | T | C | 0.22 | -0.186 | 0.009 | 3.2E-86 | 387.3 | -0.208 | 0.035 | 4.49E-09 | 1.119 | 0.199 | 1.85E-08 | 4.27E-02 | 20 |
| AOC1 | 7 | 150540153 | rs7806458 | 7 | 150476888 | G | A | 0.36 | 0.669 | 0.008 | 0.00E+00 | 6991.5 | 0.078 | 0.031 | 1.27E-02 | 0.117 | 0.047 | 1.28E-02 | 9.06E-02 | 20 |
| SCNN1D | 1 | 1221612 | rs11804831 | 1 | 1194804 | C | T | 0.15 | -0.133 | 0.012 | 4.69E-30 | 129.7 | -0.161 | 0.041 | 6.90E-05 | 1.208 | 0.321 | 1.72E-04 | 1.13E-02 | 20 |
| SLC12A1 | 15 | 48540068 | rs964611 | 15 | 48597514 | A | C | 0.15 | 1.664 | 0.009 | 0.00E+00 | 37527.1 | -0.093 | 0.042 | 2.52E-02 | -0.056 | 0.025 | 2.53E-02 | 4.70E-01 | 20 |
| CA4 | 17 | 58237778 | rs34820870 | 17 | 58244021 | G | T | 0.03 | -0.462 | 0.021 | 6.1E-110 | 496.3 | 0.202 | 0.085 | 1.74E-02 | -0.438 | 0.185 | 1.80E-02 | 1.64E-02 | 20 |
| SLC16A1 | 1 | 113477052 | rs3789592 | 1 | 113471400 | A | G | 0.41 | 0.123 | 0.008 | 4.34E-53 | 235.2 | -0.178 | 0.031 | 6.28E-09 | -1.443 | 0.266 | 5.61E-08 | 6.64E-01 | 20 |
| ADRB2 | 5 | 148207176 | rs2082395 | 5 | 148200600 | A | G | 0.42 | 0.126 | 0.008 | 6.31E-56 | 248.2 | -0.119 | 0.030 | 8.83E-05 | -0.944 | 0.248 | 1.42E-04 | 7.66E-01 | 5 |
| KCNJ11 | 11 | 17409142 | rs2074310 | 11 | 17421886 | T | C | 0.37 | 0.157 | 0.012 | 4.37E-38 | 166.5 | 0.340 | 0.032 | 5.50E-27 | 2.160 | 0.262 | 1.51E-16 | 6.53E-02 | 20 |
| ATP1A1 | 1 | 116934086 | rs6704439 | 1 | 116867616 | A | C | 0.27 | 0.041 | 0.009 | 4.39E-06 | 21.1 | 0.147 | 0.035 | 2.39E-05 | 3.567 | 1.148 | 1.88E-03 | 2.09E-01 | 8 |
| SLC9A1 | 1 | 27459389 | rs12751422 | 1 | 27493679 | T | C | 0.27 | 0.106 | 0.009 | 1.19E-32 | 141.6 | -0.148 | 0.035 | 2.08E-05 | -1.395 | 0.348 | 6.08E-05 | 9.72E-02 | 20 |
| NR3C1 | 5 | 142736286 | rs4912908 | 5 | 142791133 | G | A | 0.23 | 0.073 | 0.010 | 1.07E-13 | 55.2 | 0.094 | 0.038 | 1.20E-02 | 1.296 | 0.545 | 1.74E-02 | 2.92E-01 | 15 |
| CACNA1H | 16 | 1237506 | rs35198836 | 16 | 1244631 | T | C | 0.22 | 0.149 | 0.011 | 2.95E-39 | 171.8 | 0.099 | 0.035 | 4.71E-03 | 0.668 | 0.241 | 5.69E-03 | 3.96E-01 | 20 |
| AQP1 | 7 | 30929070 | rs28362721 | 7 | 30957702 | T | C | 0.17 | 0.086 | 0.011 | 1.81E-15 | 63.3 | 0.115 | 0.041 | 4.93E-03 | 1.345 | 0.507 | 7.97E-03 | 5.12E-01 | 20 |
| JUN | 1 | 59248125 | rs2716140 | 1 | 59472397 | C | A | 0.40 | -0.228 | 0.008 | 3.5E-177 | 805.5 | 0.126 | 0.031 | 4.63E-05 | -0.551 | 0.137 | 5.58E-05 | 9.09E-02 | 20 |
| CA12 | 15 | 63643968 | rs12909041 | 15 | 63743677 | A | C | 0.20 | 0.039 | 0.010 | 0.000114 | 14.9 | 0.090 | 0.038 | 1.92E-02 | 2.319 | 1.158 | 4.53E-02 | 1.08E-02 | 5 |
| CACNA1D | 3 | 53687586 | rs9830632 | 3 | 53735766 | G | A | 0.29 | -0.085 | 0.009 | 2E-22 | 94.9 | 0.207 | 0.034 | 9.24E-10 | -2.427 | 0.468 | 2.14E-07 | 3.93E-01 | 20 |
| CACNA2D2 | 3 | 50470954 | rs62260815 | 3 | 50474624 | A | G | 0.11 | -0.142 | 0.013 | 8.98E-29 | 123.9 | -0.116 | 0.049 | 1.67E-02 | 0.814 | 0.348 | 1.94E-02 | 7.93E-02 | 20 |
| PDE1B | 12 | 54958078 | rs1874308 | 12 | 54954356 | A | T | 0.30 | 0.429 | 0.009 | 0.00E+00 | 2301.7 | -0.094 | 0.034 | 5.33E-03 | -0.220 | 0.079 | 5.40E-03 | 7.35E-01 | 20 |
| AHR | 7 | 17362011 | rs17700436 | 7 | 17167825 | T | C | 0.05 | 0.480 | 0.017 | 1.6E-178 | 811.7 | -0.184 | 0.066 | 5.34E-03 | -0.383 | 0.138 | 5.58E-03 | 7.08E-01 | 20 |
| GPR35 | 2 | 241557762 | rs2975788 | 2 | 241571858 | G | A | 0.48 | 0.293 | 0.009 | 6.2E-259 | 1181.6 | 0.065 | 0.030 | 3.29E-02 | 0.221 | 0.104 | 3.34E-02 | 7.59E-01 | 20 |
| KCNJ2 | 17 | 68170501 | rs9890133 | 17 | 68169005 | G | A | 0.11 | -0.256 | 0.012 | 8.02E-94 | 422.2 | 0.094 | 0.047 | 4.61E-02 | -0.367 | 0.185 | 4.72E-02 | 6.24E-01 | 20 |

Abbreviations: MR, Mendelian randomization, SNP, single nuclear polymorphism, eQTL, expression quantitative trait loci, se, standard error, SBP, systolic blood pressure, HEIDI, heterogeneity in dependent instruments, nsnp, number of SNPs for HEIDI test.

**Supplementary Table 4** Association between SLC12A2 and PDE1B eQTL SNP in blood with expression of other nearby genes

| | Nearby gene | Chr | BP | Samples | eQTL SNP | SNP_BP | Effect_allele | Other_allele | Freq_Effect_allele | beta | se | p |
|---|---|---|---|---|---|---|---|---|---|---|---|---|
| SLC12A2 | **CTC-228N24.3** | **5** | **127347455** | **22495** | **rs17764730** | **127357526** | **T** | **C** | **0.227** | **-0.503** | **0.011** | **0.00E+00** |
| | **SLC12A2** | **5** | **127472419** | **31644** | **rs17764730** | **127357526** | **T** | **C** | **0.227** | **-0.186** | **0.009** | **3.20E-86** |
| | **FBN2** | **5** | **127794239** | **31684** | **rs17764730** | **127357526** | **T** | **C** | **0.227** | **-0.158** | **0.009** | **6.90E-63** |
| | PRRC1 | 5 | 126872041 | 31684 | rs17764730 | 127357526 | T | C | 0.227 | -0.019 | 0.009 | 5.01E-02 |
| | CTC-228N24.1 | 5 | 127158204 | 15590 | rs17764730 | 127357526 | T | C | 0.227 | 0.013 | 0.014 | 3.37E-01 |
| | HNRNPKP1 | 5 | 126847845 | 19056 | rs17764730 | 127357526 | T | C | 0.227 | -0.004 | 0.012 | 7.30E-01 |
| | C5orf63 | 5 | 126393717 | 25262 | rs17764730 | 127357526 | T | C | 0.227 | 0.003 | 0.011 | 7.81E-01 |
| PDE1B | **PDE1B** | **12** | **54958078** | **28131** | **rs1874308** | **54954356** | **A** | **T** | **0.289** | **0.429** | **0.009** | **0.00E+00** |
| | **NCKAP1L** | **12** | **54914610** | **28131** | **rs1874308** | **54954356** | **A** | **T** | **0.289** | **0.038** | **0.009** | **5.00E-05** |
| | **RP11-834C11.3** | **12** | **54484028** | **3827** | **rs1874308** | **54954356** | **A** | **T** | **0.289** | **-0.077** | **0.025** | **2.15E-03** |
| | **GPR84** | **12** | **54757250** | **28131** | **rs1874308** | **54954356** | **A** | **T** | **0.289** | **-0.026** | **0.009** | **4.92E-03** |
| | **SMUG1** | **12** | **54570653** | **27382** | **rs1874308** | **54954356** | **A** | **T** | **0.289** | **0.022** | **0.009** | **1.83E-02** |
| | RP11-1049A21.2 | 12 | 54937392 | 4652 | rs1874308 | 54954356 | A | T | 0.289 | -0.045 | 0.023 | 5.07E-02 |
| | RP11-968A15.8 | 12 | 54704641 | 3827 | rs1874308 | 54954356 | A | T | 0.289 | -0.047 | 0.025 | 6.18E-02 |
| | HNRNPA1 | 12 | 54677424 | 23056 | rs1874308 | 54954356 | A | T | 0.289 | -0.014 | 0.010 | 1.66E-01 |
| | NFE2 | 12 | 54690400 | 28131 | rs1874308 | 54954356 | A | T | 0.289 | 0.013 | 0.009 | 1.79E-01 |
| | RP11-968A15.2 | 12 | 54664623 | 4652 | rs1874308 | 54954356 | A | T | 0.289 | 0.031 | 0.023 | 1.82E-01 |
| | ATF7 | 12 | 53960919 | 11154 | rs1874308 | 54954356 | A | T | 0.289 | 0.019 | 0.015 | 1.97E-01 |
| | RP11-753H16.5 | 12 | 54804172 | 4018 | rs1874308 | 54954356 | A | T | 0.289 | 0.029 | 0.025 | 2.34E-01 |
| | ITGA5 | 12 | 54801144 | 28131 | rs1874308 | 54954356 | A | T | 0.289 | 0.011 | 0.009 | 2.52E-01 |
| | ZNF385A | 12 | 54773999 | 28131 | rs1874308 | 54954356 | A | T | 0.289 | -0.010 | 0.009 | 2.67E-01 |
| | RP11-834C11.4 | 12 | 54523254 | 21824 | rs1874308 | 54954356 | A | T | 0.289 | -0.011 | 0.011 | 2.86E-01 |
| | PHC1P1 | 12 | 55806448 | 4526 | rs1874308 | 54954356 | A | T | 0.289 | -0.024 | 0.023 | 3.03E-01 |
| | ATP5G2 | 12 | 54048851 | 28131 | rs1874308 | 54954356 | A | T | 0.289 | -0.009 | 0.009 | 3.60E-01 |
| | HOXC5 | 12 | 54404387 | 27306 | rs1874308 | 54954356 | A | T | 0.289 | -0.008 | 0.009 | 3.91E-01 |
| | COPZ1 | 12 | 54720309 | 28131 | rs1874308 | 54954356 | A | T | 0.289 | -0.006 | 0.009 | 5.00E-01 |
| | HOXC4 | 12 | 54430264 | 22231 | rs1874308 | 54954356 | A | T | 0.289 | 0.007 | 0.010 | 5.21E-01 |
| | RP11-834C11.7 | 12 | 54473154 | 10394 | rs1874308 | 54954356 | A | T | 0.289 | -0.009 | 0.015 | 5.55E-01 |
| | CBX5 | 12 | 54649305 | 27917 | rs1874308 | 54954356 | A | T | 0.289 | -0.004 | 0.009 | 6.70E-01 |
| | TESPA1 | 12 | 55360166 | 21824 | rs1874308 | 54954356 | A | T | 0.289 | -0.004 | 0.011 | 6.97E-01 |
| | GTSF1 | 12 | 54858560 | 11154 | rs1874308 | 54954356 | A | T | 0.289 | 0.006 | 0.015 | 7.09E-01 |
| | CALCOCO1 | 12 | 54113216 | 28131 | rs1874308 | 54954356 | A | T | 0.289 | -0.003 | 0.009 | 7.19E-01 |

Abbreviations: SNP, single nuclear polymorphism, eQTL, expression quantitative trait loci, se, standard error.

**Supplementary Table 5** MR association between expression of nearby genes of SLC12A2 and PDE1B in blood and risk of breast cancer and ER+ breast cancer

| BC type | Target gene | Nearby gene | ProbeChr | Probe bp | topSNP | topSNP_bp | Effect allele | Other allele | Freq Effect allele | eQTL association | | | BC association | | | MR association | | | HEIDI Test | | Posterior probability |
|---|---|---|---|---|---|---|---|---|---|---|---|---|---|---|---|---|---|---|---|---|---|
| | | | | | | | | | | beta | se | p | beta | se | p | beta | se | p | p_HEIDI | nsnp | |
| Any BC | SLC12A2 | CTC-228N24.3 | 5 | 127347455 | rs6888037 | 127406259 | G | T | 0.21 | -0.508 | 0.011 | 0.00E+00 | 0.027 | 0.007 | 1.09E-04 | -0.054 | 0.014 | 1.15E-04 | 1.45E-01 | 20 | 77% |
| | | SLC12A2 | 5 | 127472419 | rs17764730 | 127357526 | T | C | 0.21 | -0.186 | 0.009 | 3.20E-86 | 0.028 | 0.007 | 7.57E-05 | -0.150 | 0.039 | 1.05E-04 | 1.56E-01 | 20 | 81.50% |
| | | FBN2 | 5 | 127794239 | rs79813368 | 127921198 | A | G | 0.05 | 1.262 | 0.013 | 0.00E+00 | -0.003 | 0.011 | 7.71E-01 | -0.002 | 0.009 | 7.71E-01 | 9.98E-01 | 20 | 0.33% |
| ER+ BC | SLC12A2 | CTC-228N24.3 | 5 | 127347455 | rs6888037 | 127406259 | G | T | 0.21 | -0.508 | 0.011 | 0.00E+00 | 0.029 | 0.009 | 7.99E-04 | -0.057 | 0.017 | 8.48E-04 | 1.70E-01 | 20 | 40.40% |
| | | SLC12A2 | 5 | 127472419 | rs17764730 | 127357526 | T | C | 0.21 | -0.186 | 0.009 | 3.20E-86 | 0.029 | 0.009 | 6.96E-04 | -0.158 | 0.048 | 8.67E-04 | 1.23E-01 | 20 | 40.46% |
| | | FBN2 | 5 | 127794239 | rs79813368 | 127921198 | A | G | 0.05 | 1.262 | 0.013 | 0.00E+00 | -0.004 | 0.013 | 7.78E-01 | -0.003 | 0.011 | 7.75E-01 | 1.00E+00 | 20 | 0.42% |
| | PDE1B | PDE1B | 12 | 54958078 | rs1874308 | 54954356 | A | T | 0.39 | 0.429 | 0.009 | 0.00E+00 | 0.030 | 0.008 | 3.38E-04 | 0.070 | 0.019 | 3.28E-04 | 2.37E-01 | 14 | 66.80% |
| | | NCKAP1L | 12 | 54914610 | rs11609712 | 54890515 | A | C | 0.12 | 0.187 | 0.009 | 5.93E-100 | 0.010 | 0.009 | 2.69E-01 | 0.053 | 0.049 | 2.72E-01 | 1.03E-01 | 20 | 0.62% |
| | | GPR84 | 12 | 54757250 | rs3809161 | 54758330 | A | G | 0.50 | 0.222 | 0.008 | 3.93E-175 | -0.015 | 0.008 | 6.21E-02 | -0.067 | 0.036 | 6.16E-02 | 4.08E-01 | 20 | 1.67% |
| | | SMUG1 | 12 | 54570653 | rs2233921 | 54575800 | A | C | 0.32 | -0.344 | 0.008 | 0.00E+00 | -0.005 | 0.008 | 5.29E-01 | 0.014 | 0.022 | 5.31E-01 | 1.42E-01 | 20 | 0.34% |

Abbreviations: MR, Mendelian randomization, SNP, single nuclear polymorphism, eQTL, expression quantitative trait loci, se, standard error, BC, breast cancer, HEIDI, heterogeneity in dependent instruments, nsnp, number of SNPs for HEIDI test.

**Supplementary Table 6** MR association between drug targeted gene expression in other tissue and overall risk of breast cancer

| Tissue | Gene | Gene chr | Gene_start | topSNP | topSNP_bp | Effect allele | Other allele | Freq Effect allele | eQTL association | | | BC association | | | MR association | | | HEIDI Test | |
|---|---|---|---|---|---|---|---|---|---|---|---|---|---|---|---|---|---|---|---|
| | | | | | | | | | beta | se | p | beta | se | p | beta | se | p | p_HEIDI | nsnp |
| Adrenal Gland | KCNH2 | 7 | 150944961 | chr7_150901391 | 150901391 | C | T | 0.452 | -0.353 | 0.058 | 1.51E-09 | 0.017 | 0.010 | 1.09E-01 | -0.047 | 0.031 | 1.22E-01 | 2.49E-01 | 20 |
| | CACNA1H | 16 | 1153121 | chr16_1137931 | 1137931 | T | C | 0.408 | -0.447 | 0.061 | 3.10E-13 | -0.008 | 0.017 | 6.17E-01 | 0.019 | 0.037 | 6.18E-01 | 3.04E-01 | 12 |
| Breast Mammary | GPR35 | 2 | 240605431 | chr2_240621671 | 240621671 | T | G | 0.359 | -0.378 | 0.041 | 9.62E-21 | -0.025 | 0.014 | 7.97E-02 | 0.065 | 0.038 | 8.50E-02 | 8.58E-01 | 13 |
| | PDE1B | 12 | 54549350 | chr12_54561540 | 54561540 | C | T | 0.693 | 0.310 | 0.027 | 9.15E-30 | -0.038 | 0.011 | 6.26E-04 | -0.122 | 0.037 | 1.06E-03 | 4.72E-01 | 11 |
| Kidney Cortex | ACE | 17 | 63477061 | chr17_63482264 | 63482264 | G | A | 0.633 | 0.565 | 0.103 | 4.16E-08 | -0.008 | 0.010 | 4.28E-01 | -0.014 | 0.018 | 4.33E-01 | NA | NA |
| Muscle Skeletal | GPR35 | 2 | 240605431 | chr2_240621671 | 240621671 | T | G | 0.359 | -0.438 | 0.037 | 2.14E-32 | -0.025 | 0.014 | 7.97E-02 | 0.056 | 0.032 | 8.30E-02 | 6.89E-01 | 12 |
| | SLC12A2 | 5 | 128083766 | chr5_128146900 | 128146900 | G | A | 0.190 | 0.203 | 0.032 | 1.88E-10 | -0.017 | 0.013 | 1.98E-01 | -0.084 | 0.067 | 2.07E-01 | 6.30E-02 | 18 |
| | KCNJ11 | 11 | 17385859 | chr11_17374886 | 17374886 | C | CCTTTAAT | 0.218 | -0.152 | 0.026 | 7.82E-09 | -0.007 | 0.013 | 6.03E-01 | 0.043 | 0.083 | 6.04E-01 | 4.00E-01 | 13 |
| | ASIC1 | 12 | 50057548 | chr12_50068355 | 50068355 | C | T | 0.327 | -0.189 | 0.034 | 1.99E-08 | -0.007 | 0.012 | 5.68E-01 | 0.036 | 0.063 | 5.70E-01 | 1.45E-01 | 17 |
| Ovary | GPR35 | 2 | 240605431 | chr2_240621671 | 240621671 | T | G | 0.359 | -0.523 | 0.063 | 8.97E-17 | -0.025 | 0.014 | 7.97E-02 | 0.047 | 0.027 | 8.64E-02 | 6.79E-01 | 4 |
| | KCNH2 | 7 | 150944961 | chr7_150849357 | 150849357 | T | C | 0.033 | -1.095 | 0.189 | 6.86E-09 | -0.007 | 0.029 | 8.20E-01 | 0.006 | 0.026 | 8.20E-01 | 8.52E-01 | 16 |
| | ASIC1 | 12 | 50057548 | chr12_50089249 | 50089249 | A | G | 0.325 | -0.391 | 0.069 | 1.75E-08 | -0.009 | 0.012 | 4.32E-01 | 0.024 | 0.031 | 4.36E-01 | 2.16E-01 | 20 |
| Small Intestine Terminal Ileum | GPR35 | 2 | 240605431 | chr2_240621671 | 240621671 | T | G | 0.359 | -0.261 | 0.038 | 4.18E-12 | -0.025 | 0.014 | 7.97E-02 | 0.094 | 0.055 | 8.93E-02 | 1.61E-01 | 3 |
| Uterus | NONE | | | | | | | | | | | | | | | | | | |

**Supplementary Table 7** Causal association between systolic blood pressure (exposure) and risk of breast cancer (outcome)

| Method | Number of SNPs | beta | se | OR | 95% CI | p |
|---|---|---|---|---|---|---|
| GSMR | 521 | -0.0310 | 0.0171 | 0.969 | 0.938-1.002 | 0.0693 |
| MR Egger | 745 | -0.0095 | 0.0043 | 0.991 | 0.982-0.999 | 0.0295 |
| Weighted median | 745 | 0.0004 | 0.0017 | 1.000 | 0.997-1.004 | 0.8306 |
| Inverse variance weighted | 745 | -0.0002 | 0.0016 | 1.000 | 0.997-1.003 | 0.9178 |
| Simple mode | 745 | 0.0005 | 0.0061 | 1.000 | 0.989-1.013 | 0.9359 |
| Weighted mode | 745 | 0.0005 | 0.0038 | 1.000 | 0.993-1.008 | 0.8971 |

Abbreviations: se, standard error, SNP, single nucleotide polymorphism, OR odds ratio, CI confidence intervals

**Supplementary Table 8** Comparison of gene expression of SLC12A2 between breast cancer patients and controls in Expression Atlas

| Experiment accession | Comparison (normal as reference) | Sample part | Log_2 fold change | Adjusted p-value |
|---|---|---|---|---|
| E-GEOD-68086 | 'breast carcinoma' vs 'normal' | blood platelet | -3.3 | 7.70E-24 |
| E-GEOD-31138 | invasive ductal carcinoma' vs 'normal' | breast | 2.5 | 0.02863135 |
| E-GEOD-45581 | 'non-inflammatory breast cancer' vs 'normal' | breast | -1.5 | 0.029033235 |
| E-GEOD-54002 | 'breast cancer' vs 'normal' | mammary gland | -1.4 | 1.23364E-05 |
| E-GEOD-38959 | 'breast cancer; breast' vs 'normal; breast' | breast | -1.3 | 0.017083027 |
