## Supplemental Figure 1 for "Use of antihypertensive drugs and breast cancer risk: a two-sample Mendelian randomization study"

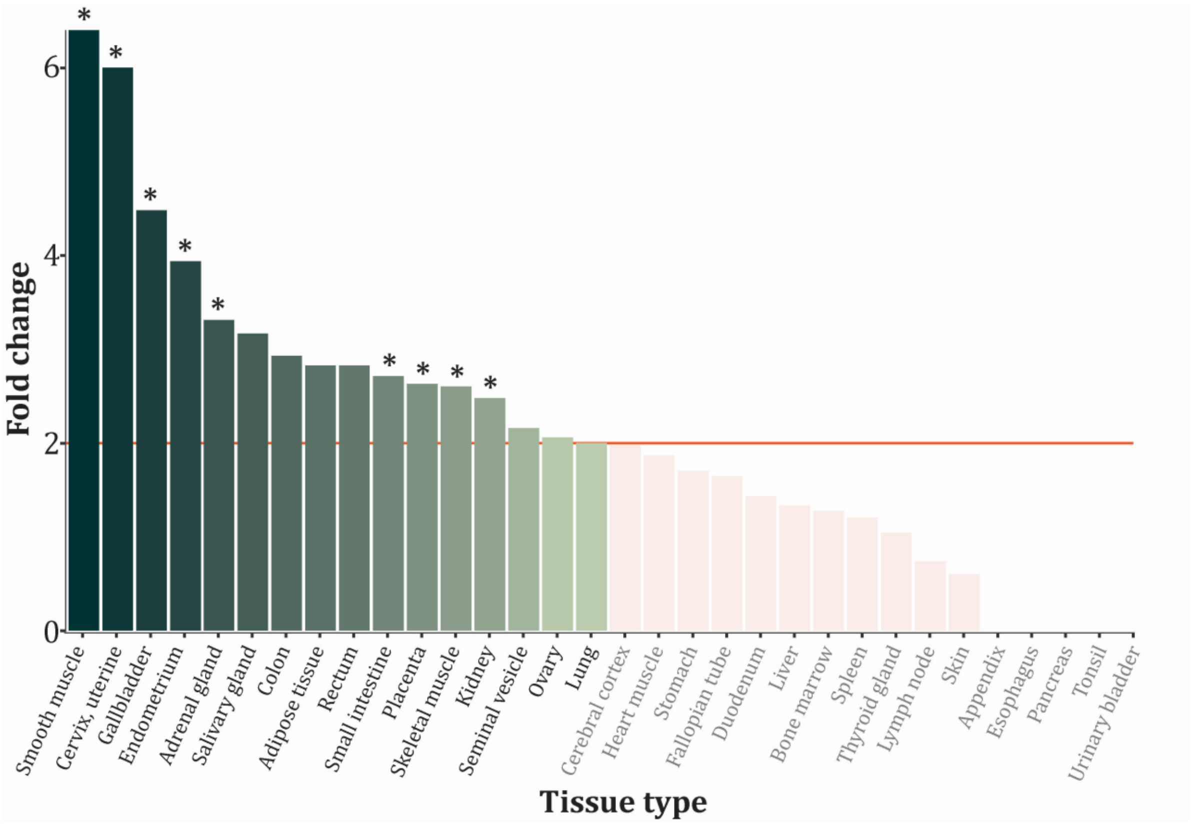


Supplementary Figure 1. Tissue enrichment identifies tissue types where the target genes are most likely to be differentially expressed. Color gradient represents decreasing fold change and light reds are insignificant associations. Stars indicate statistical significance in the tests.
